# Factors Associated with Disparities in Appropriate Statin Therapy in an Outpatient Inner City Population

**DOI:** 10.1101/2020.08.14.20174185

**Authors:** Giselle A. Suero-Abreu, Aris Karatasakis, Sana Rashid, Maciej Tysarowski, Analise Y. Douglas, Richa Patel, Emaad Siddiqui, Aishwarya Bhardwaj, Christine M. Gerula, Daniel Matassa

## Abstract

**Introduction:** Appropriate lipid-lowering therapies are essential for the primary and secondary prevention of atherosclerotic cardiovascular disease (ASCVD). The aim of this study is to identify discrepancies between cholesterol management guidelines and current practice in an underserved population, with a focus on statin treatment.

**Methods:** We reviewed the records of 1,042 consecutive patients seen between August 2018 and August 2019 in an outpatient academic primary care clinic. Eligibility for statin and other lipid-lowering therapies was determined based on the 2018 American Heart Association and American College of Cardiology (AHA/ACC) guideline on the management of blood cholesterol.

**Results:** Among 464 statin-eligible patients, age was 61.1 ± 10.4 years and 53.9% were female. Most patients were Black (47.2%), followed by Hispanic (45.7%), and White (5.0%). Overall, 82.1% of patients were prescribed a statin. Statin-eligible patients who qualified based only on a 10-year ASCVD risk > 7.5% were less likely to be prescribed a statin (32.8%, p<0.001). After adjustment for gender and health insurance status, appropriate statin treatment was associated with age > 55 years (OR = 4.59 [95% CI 1.09 - 16.66], p = 0.026), hypertension (OR = 2.38 [95% CI 1.29 - 4.38], p = 0.005) and chronic kidney disease (OR = 3.95 [95% CI 1.42 - 14.30], p = 0.017). Factors independently associated with statin undertreatment were Black race (OR = 0.42 [95% CI 0.23 - 0.77], p = 0.005), and statin-eligibility based solely on an elevated 10-year ASCVD risk (OR = 0.14 [95% CI 0.07 - 0.25], p < 0.001). Hispanic patients were more likely to be on appropriate statin therapy when compared to Black patients (86.8% vs 77.2%).

**Conclusion:** Statin underprescription is seen in approximately one out of five eligible patients, and is independently associated with Black race, younger age, fewer comorbidities, and eligibility via 10-year ASCVD risk only. Hispanic patients are more likely to be on appropriate statin therapy compared to Black patients.

## Introduction

Atherosclerotic cardiovascular disease (ASCVD) is a leading cause of morbidity and mortality globally, with over 600,000 deaths in the United States annually.^1,2^ Statins are an effective therapy for the primary and secondary prevention of ASCVD with proven mortality benefits and robust long-term safety data.^3^ Reflecting this, current ASCVD prevention guidelines including the American Heart Association and American College of Cardiology (AHA/ACC) multisociety guidelines on the management of blood cholesterol have recommended statins as the cornerstone of lipid-lowering therapy (LLT) for primary and secondary ASCVD prevention.^4^ Nonetheless, gaps exist in guideline-directed statin prescription patterns in clinical practice. For example, racial and ethnic minorities and patients with socioeconomic barriers to healthcare face disparities that result in worse outcomes and higher mortality rates secondary to ASCVD.^5^ Younger Black patients are particularly vulnerable to these disparities in statin prescription patterns.^6^

This effect of age and race on statin prescription compounded by a general delay in the adoption of new guidelines by healthcare providers, leads to suboptimal patient care in these populations.^7^ The objectives of this study were 1) to identify discrepancies between cholesterol management guidelines and current practice in an inner city academic center primary care population; and 2) to provide insights into possible interventions aimed at improving statin prescribing patterns and thus reducing the incidence of ASCVD in this vulnerable cohort.

## Methods

We retrospectively analyzed 1,042 consecutive patient encounters taking place between August 2018 and August 2019 at the University Hospital Ambulatory Care Center, Newark, NJ, and determined eligibility for statins and other LLT in adults aged 20-75 years based on the 2018 AHA/ACC multisociety guideline on the management of blood cholesterol. We identified 509 statin-eligible patients, out of which 464 patients were included in the final analysis; 45 patients met the exclusion criterion of well-documented contraindications to statins (e.g., allergy, patient preference, or side effects) and were removed from the analysis (Figure 1). High-intensity statins were defined as Atorvastatin 40mg - 80mg and Rosuvastatin 20mg - 40mg. All data was obtained from the electronic health record and this study was approved by the Institutional Review Board. The study met the requirements of the Declaration of Helsinki and was performed in compliance with human-studies guidelines. Individual consent for participation in anonymous data analysis was waived. Data was collected and managed via Research Electronic Data Capture (REDCap), a secure web-based software platform designed to support data capture hosted at Rutgers New Jersey Medical School.^8^ The pooled cohort equation was used to estimate ASCVD risk and criteria for statin-eligibility was determined based on guidelines current at the time of the patient encounter.4 Data is presented as mean ± standard deviation for normally distributed continuous variables or median (interquartile range) for non-normally distributed continuous variables, and compared using the t-test or the Wilcoxon rank-sum test, respectively. Categorical variables are presented as a number (percentage) and compared using the chi-square as appropriate. Multivariate logistic regression analysis was performed to identify independent predictors of statin prescription in the cohort; variables with a p < 0.2 were entered into a backward elimination procedure, along with gender and health insurance status. Odds ratios (OR) were calculated to determine the association between patient characteristics and statin treatment status. All statistical analyses were two-sided, and significance was established at α = 0.05. Analyses were performed using R statistical software version 3.6.1 R Foundation for Statistical Computing, Vienna, Austria.

**Figure 1.**
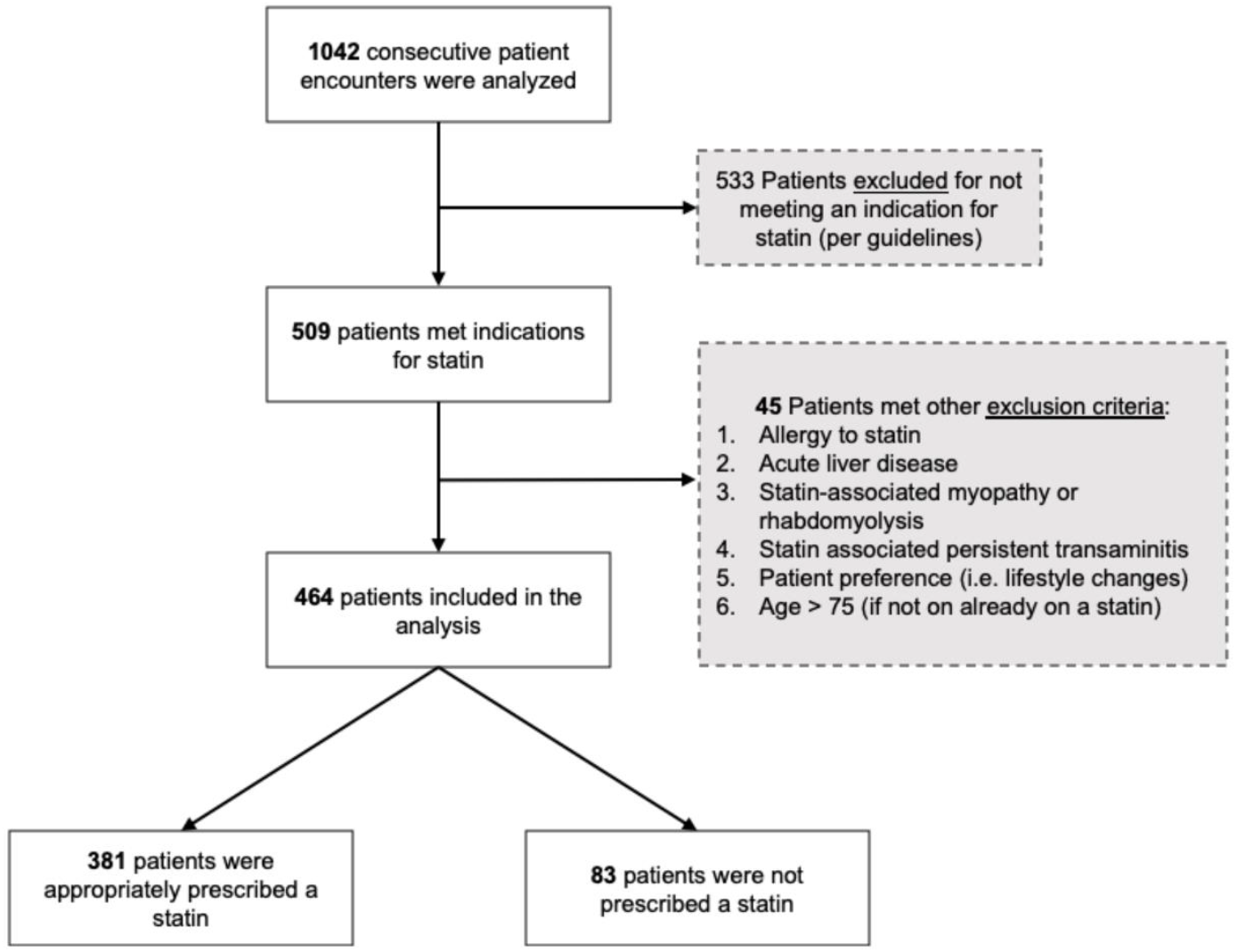
Flow diagram demonstrating subject inclusion and exclusion criteria and patients that were prescribed a statin.

## Results

Among 464 statin-eligible patients, the average age was 61.1 ± 10.43 and the majority were female (53.9%). Additionally, patients who identified as Black (47.2%) and Hispanic or Latino (45.7%) comprised the greatest number of patients, followed by White (5.0%), Asian (1.9%), and Pacific Islander (0.2%). Furthermore, 38.8% did not report English as their primary language, out of which 70.6% needed translation services during the encounter. Most of the patients lacked health insurance (54.7%). The most common comorbidities were hypertension (HTN, 81.0%), diabetes mellitus (DM, 45.0%), chronic kidney disease (CKD, 15.3%), cerebrovascular disease (CVD, 14.7%), coronary artery disease (CAD, 14.6%), heart failure (HF, 12.9%), and peripheral artery disease (PAD, 2.8%). Additionally, 17.9% of patients were current smokers. A cardiologist was seen within 12 months of the index encounter for 30.4% (n=141) of patients, out of which 92.2% were placed on an appropriate statin. A lipid profile was available for 97.4% (n=452) of patients and the median LDL-C was 87 mg/dl [IQR 64.0 - 120.0]. Among diabetic patients, the median HbA1c was 7.9% [IQR 6.5, 8.9]. Of the 464 statin-eligible patients, statin benefit groups were defined based on 2018 AHA/ACC guidelines as: very high-risk ASCVD (16.4% n= 76), clinical ASCVD (10.3% n=48), 10-year ASCVD risk ≥7.5% (39.9% n=185), DM in 40-75 years old (31.3% n=145), and LDL-C > 190mg/dl (2.2% n=10) and the rate of statin prescription per benefit group is shown in Figure 2. In the statin-eligible cohort (n=464), 82.1% of patients were prescribed a statin. A high-intensity statin was indicated in 69.6% of patients and was appropriately prescribed in 88.5% of these patients. Appropriate statin prescription was associated with the presence of common comorbidities such as DM, CKD, HF, CAD, CVD, and lower diastolic blood pressure (Table 1, p < 0.05). The most prescribed statins and doses were: Atorvastatin 40-80 mg (61.2%), Atorvastatin 10-20 mg (9.7%), Pravastatin 40-80 mg (4.7%), Simvastatin 20-40 mg (4.5%), and Rosuvastatin 20-40mg (0.5%). Of those already prescribed a statin, only 27.2% of patients were monitored with subsequent LDL-C ordered for treatment efficacy. Out of the 126 patients on statins who had inadequate LDL-C reduction while on the maximally tolerated statin, 40.4% met criteria for Ezetimibe and 8.7% met criteria for PCSK9 inhibitors. Persistent hypertriglyceridemia was identified in 3.2% of patients on high-intensity statins and only 0.02% were on a triglyceride-lowering therapy. There was no difference in LLT prescription related to the experience of the prescriber.

**Table 1.** Group characteristics

|  | Total<br>(n = 464) | Statin prescribed<br>(n = 381) | Statin not Prescribed<br>(n = 83) | P |
| --- | --- | --- | --- | --- |
| **DEMOGRAPHICS** |  |  |  |  |
| Age, years | 61.0 (10.4) | 61.8 (10.5) | 57.2 (9.1) | **0.0001** |
| Males, n (%) | 214 (46.1) | 177 (46.5) | 37 (44.6) |  |
| **Race/Ethnicity, n (%)** |  |  |  | 0.12 |
| Black or African American | 219 (47.2) | 169 (44.4) | 50 (60.2) |  |
| Hispanic or Latino | 212 (45.7) | 184 (48.3) | 28 (33.7) |  |
| White | 23 (5.0) | 19 (5.0) | 4 (4.8) |  |
| Asian | 9 (1.9) | 8 (2.1) | 1 (1.2) |  |
| Native Hawaiian/Pacific Islander | 1 (0.2) | 1 (0.3) | 0 (0.0) |  |
| **Primary Language, n (%)** |  |  |  | **0.500** |
| English | 284 (61.2) | 227 (59.6) | 57 (68.7) |  |
| Spanish | 130 (28.0) | 111 (29.1) | 19 (22.9) |  |
| Portuguese | 31 (6.7) | 26 (6.8) | 5 (6.0) |  |
| Other | 15 (3.2) | 14 (3.7) | 1 (1.2) |  |
| French Creole | 4 (0.9) | 3 (0.8) | 1 (1.2) |  |
| Translator utilized/Physician Fluent | 127 (71.3) | 107 (70.4) | 20 (76.9) | 0.656 |
| Uninsured, n (%) | 254 (54.7) | 207 (54.3) | 47 (56.6) | 0.796 |
| BMI, kg/m<sup>2</sup> | 29.4 [25.7, 33.4] | 29.4 [25.8, 33.3] | 29.0 [25.2, 35.1] | 0.875 |
| **COMORBIDITIES, n (%)** |  |  |  |  |
| Hypertension | 376 (81.0) | 320 (84.0) | 56 (67.5) | **0.001** |
| Systolic BP | 142.0 (65.3) | 142.1 (71.5) | 141.7 (21.0) | 0.956 |
| Diastolic BP | 73.7 (12.0) | 73.0 (11.5) | 77.0 (13.5) | **0.006** |
| Diabetes Mellitus | 209 (45.0) | 209 (45.0) | 209 (45.0) | **0.0001** |
| Hemoglobin A1c | 7.2 [6.5, 8.9] | 7.2 [6.5, 8.9] | 7.2 [6.5, 8.9] | 0.063 |
| Chronic kidney disease | 71 (15.3) | 67 (17.6) | 4 (4.8) | **0.006** |
| Congestive heart failure | 60 (12.9) | 56 (14.7) | 4 (4.8) | **0.024** |
| Coronary artery disease | 67 (14.4) | 67 (17.6) | 0 (0.0) | **0.0001** |
| History of MI | 32 (6.9) | 31 (8.2) | 1 (1.2) | **0.043** |
| History of PCI | 30 (6.5) | 30 (7.9) | 0 (0.0) | **0.017** |
| History of CABG | 15 (3.2) | 15 (3.9) | 0 (0.0) | 0.135 |
| Cerebrovascular disease | 68 (14.7) | 65 (17.1) | 3 (3.6) | **0.003** |
| Peripheral Artery Disease | 13 (2.8) | 12 (3.1) | 1 (1.2) | 0.545 |
| Current Smoker | 83 (17.9) | 63 (16.5) | 20 (24.1) | 0.206 |
| Lipid Profile, mg/dL |  |  |  |  |
| LDL-C | 87.0 [64.0, 120.0] | 84.0 [62.5, 121.0] | 93.0 [72.0, 112.5] | 0.339 |
| HDL-C | 50.0 [41.0, 59.0] | 49.0 [40.0, 58.0] | 52.0 [43.8, 63.8] | **0.048** |
| **Triglycerides** | 103.0 [78.0, 149.0] | 104.5 [80.2, 150.0] | 95.5 [69.5, 135.8] | 0.056 |
| **STATIN INDICATIONS, n (%)** |  |  |  | **0.0001** |
| (A) Very high-risk ASCVD | 76 (16.4) | 73 (19.2) | 3 (3.6) |  |
| (B) Clinical ASCVD | 48 (10.3) | 46 (12.1) | 2 (2.4) |  |
| (C) 10-year ASCVD risk >7.5 | 185 (39.9) | 125 (32.8) | 60 (72.3) |  |
| (D) Diabetes, age 40- 75 | 145 (31.2) | 127 (33.3) | 18 (21.7) |  |
| (E) LDL-C >190mg/dL | 10 (2.2) | 10 (2.6) | 0 (0.0) |  |

Values represent mean ± standard deviation, median [IQR; 25th –75th percentiles], or number (%). Bold values indicate statistical significance (P<0.05). CABG = coronary artery bypass graft; HDL-C = high-density lipoprotein cholesterol; LDL-C = low-density lipoprotein cholesterol; MI = myocardial infarction; PCI = percutaneous coronary intervention.

Of patients not on statins in the statin-eligible cohort (17.9%, n = 83), the majority were female (55.4%). Furthermore, 60.2% were Black, and patients were relatively younger than those on appropriate statin therapy (57.2 years old ± 9.1 vs 61.8 ± 10.5, p = 0.0001). Interestingly, 72.3% of statin-eligible patients not placed on statins, met the indication for statin solely based on a calculated 10-year ASCVD of > 7.5% (p < 0.0001). Among this group, 32.4% did not have a statin prescribed; this was therefore the set of patients with the lowest prescribing rate among all statin benefit groups. In a multivariate analysis after adjustment for gender and insurance status, appropriate statin treatment correlated positively with older age (OR = 4.59 [95% CI 1.09 - 16.66], p = 0.026), HTN (OR = 2.38 [95% CI 1.29 - 4.38], p = 0.005) and CKD (OR = 3.95 [95% CI 1.42 - 14.30], p = 0.017). Race was a significant predictor of statin prescribing and Black patients were less likely to receive a statin (OR = 0.42 [95% CI 0.23 - 0.77], p = 0.005). The negative correlation was similar for the 10-year ASCVD risk >7.5% only benefit group (OR = 0.14 [95% CI 0.07 - 0.25], p < 0.001) (Figure 2). Conversely, Hispanic patients were more likely to be on appropriate statin therapy when compared to Black patients (86.8% vs 77.2%). There was no association between appropriate statin therapy and health insurance status or the gender of the patient in our stepwise logistic regression model.

**Figure 2.**
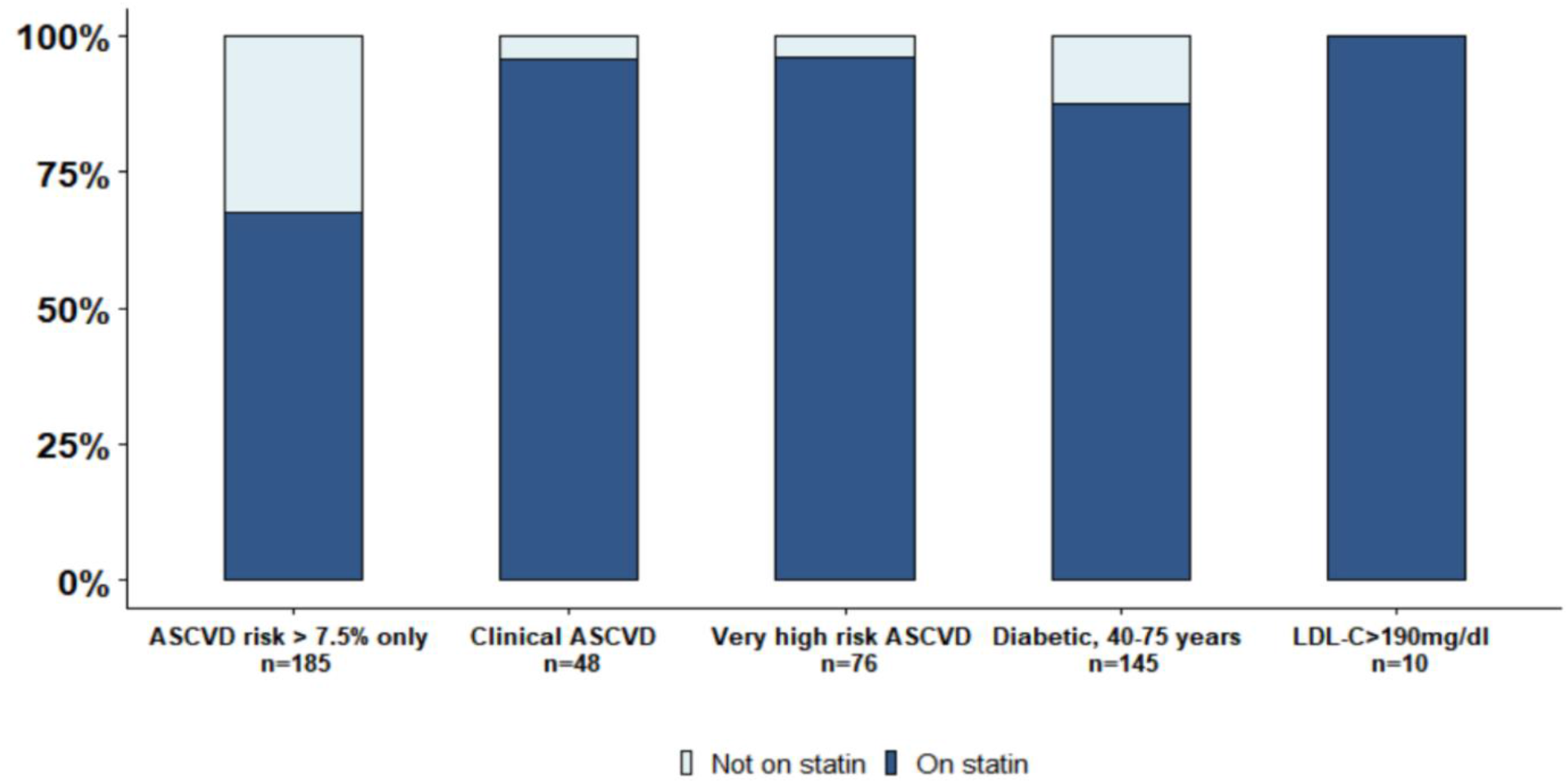
Rate of statin prescription and underprescription based on eligibility group

## Discussion

The AHA/ACC cholesterol guidelines emphasize appropriate statin therapy and the monitoring of its efficacy in statin-eligible groups to reduce the risk of micro- and macrovascular complications related to dyslipidemia. Our study demonstrated that the majority of statin-eligible patients were prescribed a statin following best prescribing patterns for secondary prevention groups (e.g., very high- risk ASCVD, clinical ASCVD). However, most patients did not have subsequent LDL-C measurements to assess the effectiveness of statin therapy. This may be associated with some patients not being maximized on statin therapy and could have, thus, contributed to lower prescription of adjunct LLTs, such as ezetimibe and PCSK-9 inhibitors.

In addition, younger patients were more likely to meet statin indication solely with 10-year ASCVD risk assessment of ≥ 7.5% and were also less likely to be prescribed a statin. In this context, underutilization of the 10-year ASCVD risk calculator could have led to the underpresciption of statins in younger patients with fewer comorbidities who met statin eligibility solely with an ASCVD risk of ≥ 7.5%. This shows that assessment of the 10-year ASCVD risk is paramount to early primary prevention in younger patients and that there is a need to re-stratify these patients based on risk enhancers according to the 2018 ACC/AHA guidelines. It is also noteworthy that since ASCVD manifests as early as the fifth decade of life, and given the proven mortality benefit of statins, early primary prevention is crucial to prevent CVD events and death, with each 1mg/dL LDL-C reduction correlating to a 1% decrease in the risk of CVD.^3,4,9^

**Figure 3.**
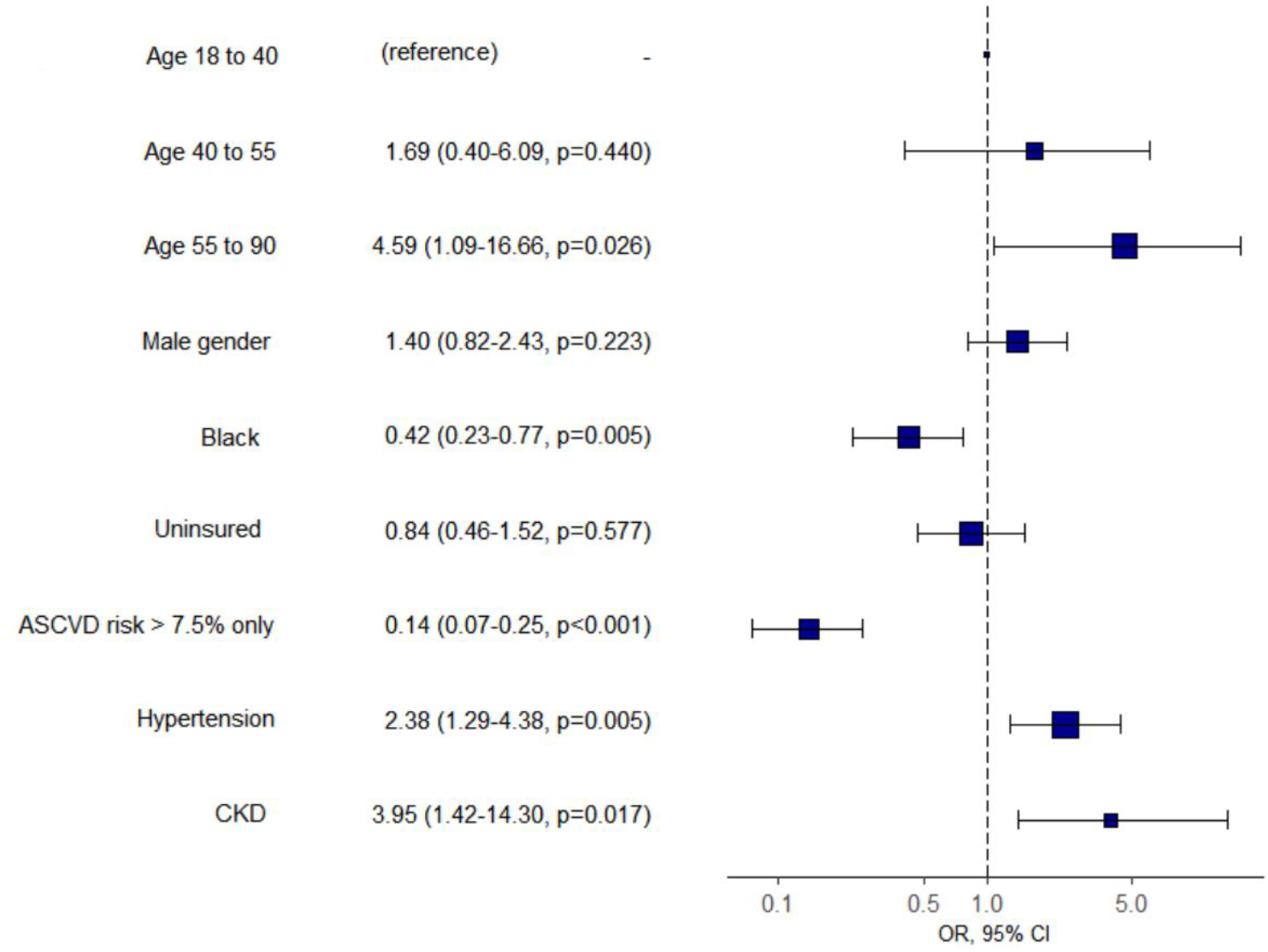
Multivariate analysis: appropriate statin prescription (95% CI, p- value)

In our study, most of the patient population was either Black or Hispanic with lower statin prescription rates noted in Black patients. In particular, younger Black patients were less likely to be on an appropriate statin, especially when the sole indication was a 10-year ASCVD risk score of ≥7.5%. Health disparities in Black patients with CVD have previously been demonstrated in several studies.^6,10,11^ Dorsch et al conducted a retrospective study in 2019 with over 9000 participants which showed statin underprescription in younger Black patients when compared with majority White patients. Similarly, the PALM (Patient and Provider Assessment of Lipid Management) registry found that Black patients were underprescribed statins across multiple specialties when compared to White patients and the REGARDS (Reasons for Geographic and Racial Differences in Stroke) study cited lower rates of statin use by Black patients living in poverty who lacked health insurance.^11,12^ Our study expands on this data by comparing Black patients to a largely Hispanic population. Although CVD is the leading cause of death among Hispanic patients, they remain underrepresented in studies.^13^ When compared with Black patients in our study, Hispanics had a higher statin prescription rate. The wide array of ethnic groups with distinct genetic predispositions and socioeconomic backgrounds amongst the Hispanic population, makes it challenging to draw overarching conclusions. A study of over 16,000 Hispanic patients confirmed an association between lower socioeconomic class, Spanish language preference, and other cardiovascular risk factors with higher rates of dyslipidemia.^13^ Furthermore, Cubans and South Americans tend to have higher rates of dyslipidemia, while Dominicans and Puerto Ricans have higher statin adherence and awareness of their hyperlipidemia.^13-15^

Statin underprescription is complex and a multifaceted approach in the comprehension of these barriers is needed to address this healthcare burden. Physician bias, low health literacy, poor follow-up, high rate of problem-based versus preventative visits, and language barriers account for some of these challenges. Although our data is limited to a single center, this study reflects disparities in statin prescription patterns in a population where most patients represent minority communities. Our data offers a unique insight on statin prescription patterns whereby even in a minority underserved setting, Black patients continue to suffer from undertreatment.

## Conclusion

The primary and secondary prevention of ASCVD is dependent on optimal guideline-directed pharmacotherapy. The AHA/ACC cholesterol guidelines emphasize proper screening and treatment monitoring to ensure that lipid-lowering agents are appropriately dosed and adjusted. Our population of patients provides a unique insight into healthcare disparities in statin prescription patterns. Although our center has a plurality of Black patients, our data has similarities to other studies where Black patients did not comprise such a large percentage of the population and also provides interesting results on statin prescription patterns in Hispanics patients relative to Black patients. Our results suggest that certain known socioeconomic health disparities fail to correct when Black patients are no longer a minority group relative to other races. Because multiple systemic barriers exist to providing optimal care, comprehensive assessment of barriers to appropriate statin prescription particularly in minorities and underserved populations is paramount. In addition, emphasis needs to be placed on cardiovascular risk stratification in younger patients based on the presence of risk-enhancers and close LDL-C monitoring to evaluate treatment adherence and efficacy. Moving forward in order to improve statin prescription patterns and ultimately better cardiovascular outcomes in minority Black and Hispanic populations, we need larger studies to systematically understand the specific challenges surrounding these communities.

## Data Availability

Not available

## Author Contributions and Notes

G.A.S-A: conceptualization, methodology, investigation, validation, data curation, writing - original draft, review & editing, visualization, project administration, formal analysis, software, resources. A.K: conceptualization, methodology, investigation, validation, data curation, writing - original draft, review & editing, visualization, project administration, formal analysis, software, resources. S.R: investigation, validation, data curation, writing - original draft, review & editing, visualization, project administration, formal analysis, software, resources. M.T: validation, data curation, writing - original draft, review & editing, visualization, project administration, formal analysis, software, resources. A.D: conceptualization, methodology, investigation, review & editing. R.P: investigation, validation, review & editing; E.S: investigation, validation, review & editing; A.B: validation, review & editing; C.G: investigation, validation, review & editing; D.M: conceptualization, methodology, data curation, review & editing, visualization, project administration, formal analysis, software, resources.

## Acknowledgments

None

## Conflicts of Interest / Funding

None

## Notes

### Competing Interest Statement

The authors have declared no competing interest.

### Funding Statement

No Funding

### Author Declarations

Study was approved by the IRB of Rutgers University New Jersey Medical School

## References

1. Heron M. Deaths: Leading Causes for 2017. Natl Vital Stat Rep. 2019;68(6):1-77

2. Benjamin EJ, Muntner P, Alonso A, et al. Heart Disease and Stroke Statistics-2019 Update: A Report From the American Heart Association [published correction appears in Circulation. 2020 Jan 14;141(2):e33]. Circulation. 2019;139(10):e56-e528. doi:10.1161/CIR.0000000000000659

3. Cholesterol Treatment Trialists' (CTT) Collaborators, Mihaylova B, Emberson J, et al. The effects of lowering LDL cholesterol with statin therapy in people at low risk of vascular disease: meta-analysis of individual data from 27 randomised trials. Lancet. 2012;380(9841):581-590. doi:10.1016/S0140-6736(12)60367-5

4. Grundy SM, Stone NJ, Bailey AL, et al. 2018 AHA/ACC/AACVPR/AAPA/ABC/ACPM/ADA/AGS/APh A/ASPC/NLA/PCNA Guideline on the Management of Blood Cholesterol: A Report of the American College of Cardiology/American Heart Association Task Force on Clinical Practice Guidelines [published correction appears in Circulation. 2019 Jun 18;139(25):e1182-e1186]. Circulation. 2019;139(25):e1082-e1143. doi:10.1161/CIR.0000000000000625

5. Graham G. Disparities in cardiovascular disease risk in the United States. Curr Cardiol Rev. 2015;11(3):238-245. doi:10.2174/1573403x11666141122220003

6. Dorsch MP, Lester CA, Ding Y, Joseph M, Brook RD. Effects of Race on Statin Prescribing for Primary Prevention With High Atherosclerotic Cardiovascular Disease Risk in a Large Healthcare System. J Am Heart Assoc. 2019;8(22):e014709. doi:10.1161/JAHA.119.014709

7. Vander Schaaf EB, Seashore CJ, Randolph GD. Translating Clinical Guidelines Into Practice: Challenges and Opportunities in a Dynamic Health Care Environment. N C Med J. 2015;76(4):230-234. doi:10.18043/ncm.76.4.230

8. Harris PA, Taylor R, Thielke R, Payne J, Gonzalez N, Conde JG. Research electronic data capture (REDCap)-- a metadata-driven methodology and workflow process for providing translational research informatics support. J Biomed Inform. 2009;42(2):377-381. doi:10.1016/j.jbi.2008.08.010

9. López-Melgar B, Fernández-Friera L, Oliva B, et al. Short-Term Progression of Multiterritorial Subclinical Atherosclerosis. J Am Coll Cardiol. 2020;75(14):1617-1627. doi:10.1016/j.jacc.2020.02.026

10. Hozawa A, Folsom AR, Sharrett AR, Chambless LE. Absolute and attributable risks of cardiovascular disease incidence in relation to optimal and borderline risk factors: comparison of African American with white subjects-- Atherosclerosis Risk in Communities Study. Arch Intern Med. 2007;167(6):573-579. doi:10.1001/archinte.167.6.573

11. Schroff P, Gamboa CM, Durant RW, Oikeh A, Richman JS, Safford MM. Vulnerabilities to Health Disparities and Statin Use in the REGARDS (Reasons for Geographic and Racial Differences in Stroke) Study. J Am Heart Assoc. 2017;6(9):e005449. Published 2017 Aug 28. doi:10.1161/JAHA.116.005449

12. Nanna MG, Navar AM, Zakroysky P, et al. Association of Patient Perceptions of Cardiovascular Risk and Beliefs on Statin Drugs With Racial Differences in Statin Use: Insights From the Patient and Provider Assessment of Lipid Management Registry. JAMA Cardiol. 2018;3(8):739-748. doi:10.1001/jamacardio.2018.1511

13. Rodriguez CJ, Daviglus ML, Swett K, et al. Dyslipidemia patterns among Hispanics/Latinos of diverse background in the United States. Am J Med. 2014;127(12):1186-94.e1. doi:10.1016/j.amjmed.2014.07.026

14. Rodriguez CJ, Cai J, Swett K, et al. High Cholesterol Awareness, Treatment, and Control Among Hispanic/Latinos: Results From the Hispanic Community Health Study/Study of Latinos. J Am Heart Assoc. 2015;4(7):e001867. Published 2015 Jun 24. doi:10.1161/JAHA.115.001867

15. Qato DM, Lee TA, Durazo-Arvizu R, et al. Statin and Aspirin Use Among Hispanic and Latino Adults at High Cardiovascular Risk: Findings From the Hispanic Community Health Study/Study of Latinos. J Am Heart Assoc. 2016;5(4):e002905. Published 2016 Mar 30. doi:10.1161/JAHA.115.002905

